# Quality of Life and Psychiatric Comorbidities Associated with Migraines in Medical Students: A Cross-Sectional Study

**DOI:** 10.1101/2024.09.05.24312634

**Authors:** Nusaiba Hassan, Roaa Hassan, Batoul Mohammed, Leina Elomeiri

## Abstract

**Objectives:** This study aims to assess the health-related quality of life (HRQoL) and psychiatric comorbidities among medical students suffering from migraines at the University of Khartoum.

**Materials and Methods:** A descriptive observational cross-sectional study was conducted on 78 medical students diagnosed with migraines. Participants completed a self-reported questionnaire that included the SF-12 Health Survey to assess HRQoL and the Hopkins Symptom Checklist (HSCL-25) to evaluate psychiatric symptoms.

**Results:** 42.3% of students reported poor quality of life, particularly in the mental component (MCS) compared to the physical component (PCS). Psychiatric symptoms, including anxiety and depression, were prevalent in 70.5% of the participants. Gender was the only sociodemographic factor significantly affecting both HRQoL and psychiatric comorbidity, with females reporting poorer physical health and higher rates of psychiatric symptoms than males, consistent with findings from other studies.

**Conclusion:** Quality of life in regard to the mental aspect is significantly impaired in medical students suffering from migraine. Psychiatric comorbidities, namely depression and anxiety, are common in these patients. Universities should keep reports of students who are diagnosed with migraine and provide support programs and follow-up regarding medical treatment and psychiatric evaluation.

## Introduction

Migraines are a prevalent and debilitating neurological disorder that affects approximately 14% of the global population [1], with a higher incidence observed among young adults, including university students [2,3,4]. Findings from the 2019 Global Burden of Disease study (GBD) indicate that migraine is the second leading cause of global disability, and the first in young women [5]. For medical students, who often face high levels of academic pressure and stress, the burden of migraines can be particularly pronounced, affecting their academic performance and overall well-being [6,7].

The assessment of health-related quality of life (HRQoL) has emerged as a critical component in understanding the broader impact of chronic conditions like migraines. HRQoL encompasses both the physical and mental health domains, providing insight into how conditions like migraines affect daily functioning and psychological well-being. In addition to the physical pain and discomfort associated with migraines, there is a growing body of evidence linking migraines to psychiatric comorbidities, such as anxiety and depression [8,9]. These comorbidities can exacerbate the overall burden of migraines, leading to a vicious cycle of worsening symptoms and impaired quality of life. In addition, studies have shown that the presence of psychiatric conditions is a risk factor for the transformation of migraine into a chronic form [10].

While there is extensive research on migraines and their impact on the general population, studies focusing specifically on medical students, particularly in regions like Sudan, remain scarce. Medical students represent a unique population due to their exposure to high levels of stress, irregular sleep patterns, and the demanding nature of their studies, all of which may contribute to the onset or exacerbation of migraines. Understanding the interplay between migraines, psychiatric comorbidities, and quality of life in this population is crucial for developing targeted interventions and support systems.

This study aims to fill this gap by assessing the health-related quality of life and psychiatric comorbidities among medical students at the University of Khartoum who suffer from migraines. By exploring these associations, we hope to contribute to a better understanding of the specific challenges faced by this population and to inform strategies for improving their academic and personal well-being.

## Materials and Methods

### Study Design

This is a descriptive observational cross-sectional study conducted among medical students at the Faculty of Medicine, University of Khartoum. The study was carried out over one month in 2019.

### Sampling

Sample size was calculated from the formula:

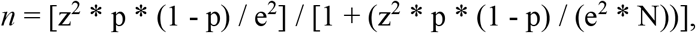

Where:

- *n* = Sample size
- *Z* = Given *Z* value = 1.96 for a confidence level of 95%
- *p* = Prevalence = 5.4%, according to the most recent findings from Alay et al. (2019) [11]
- *e* = Margin of Error = 5%
- *Pop* = Population = a total of 2111 students

Using the above equation, the sample size was calculated as 76 students.

### Participants

Inclusion criteria included students who were enrolled in the Faculty of Medicine at the University of Khartoum and had a clinical diagnosis of migraine. Exclusion criteria included students with other chronic neurological conditions or psychiatric disorders diagnosed prior to the onset of migraine. Participants were recruited through voluntary participation following an invitation sent through student email lists and announcements in classes. A total of 78 students completed the questionnaire.

### Data Collection Instruments

Data were collected using a self-administered questionnaire that included the SF-12 Health Survey and the Hopkins Symptom Checklist-25 (HSCL-25).

- **SF-12 Health Survey**: The SF-12 is a validated tool designed to reduce respondent burden while achieving minimum standards of precision for group comparisons across multiple health dimensions. It comprises eight domains: general health, physical functioning, role limitations due to physical health, role limitations due to emotional problems, bodily pain, vitality, mental health, and social functioning. These domains contribute to two summary scores: the Physical Component Summary (PCS) and the Mental Component Summary (MCS).
- **Hopkins Symptom Checklist-25 (HSCL-25)**: The HSCL-25 is a widely used instrument for assessing symptoms of anxiety and depression. It consists of two subscales: 10 items related to anxiety and 15 items related to depression. Each item is rated on a 4-point Likert scale ranging from 1 (“Not at all”) to 4 (“Extremely”). A total score is calculated by summing all 25 items, with higher scores indicating greater psychological distress. An average score of >1.75 for either anxiety or depression, or the total score across all 25 symptoms, is indicative of clinically significant symptoms. The HSCL-25 has been validated against several diagnostic instruments, including the Structured Clinical Interview for DSM-IV (SCID-IV) (Kaaya et al., 2003) [12], the Composite International Diagnostic Interview for ICD-9 (CIDI I) (Sandanger et al., 1998) [13] and ICD-10 (CIDI II) (Sandanger et al., 1999) [14], and the Present State Examination (PSE 9) (Nettelbladt et al., 1993) [15].

### Procedure

Participants were provided with an informed consent form prior to participation and the questionnaire was distributed in electronic format. Data collection was conducted anonymously to protect participants’ privacy. Ethical approval for the study was obtained from the Ethics Committee at the University of Khartoum, Faculty of Medicine, Department of Community Medicine.

### Statistical Analysis

Data were analyzed using the SPSS v.23 system. Descriptive statistics were used to summarize the demographic characteristics of the participants, as well as the SF-12 and HSCL-25 scores. Inferential Statistics were done using Pearson correlation and linear regression. A p-value of less than 0.05 was considered statistically significant.

## Results

### Participant Demographics

78 medical students participated, with a majority being female (74.4%). The mean age was 21.5 years old, ranging from 17 to 26 years old [Table 1].

**Table 1:**
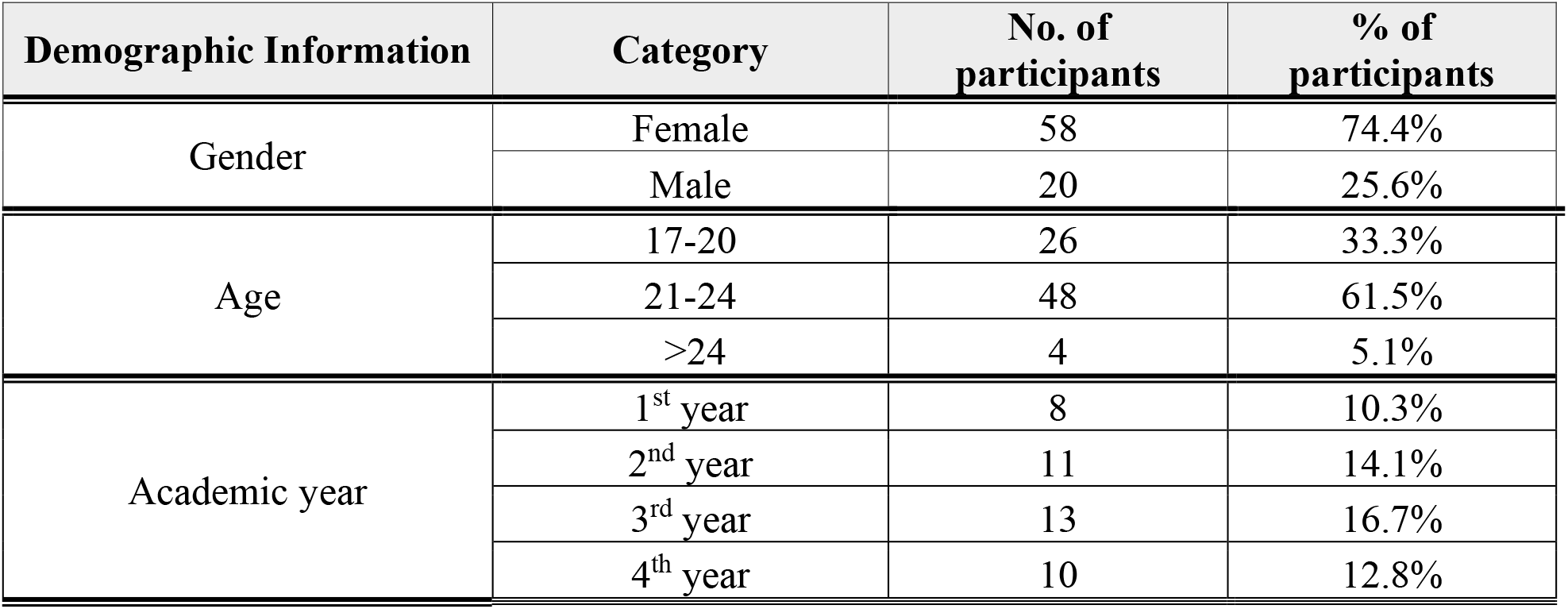
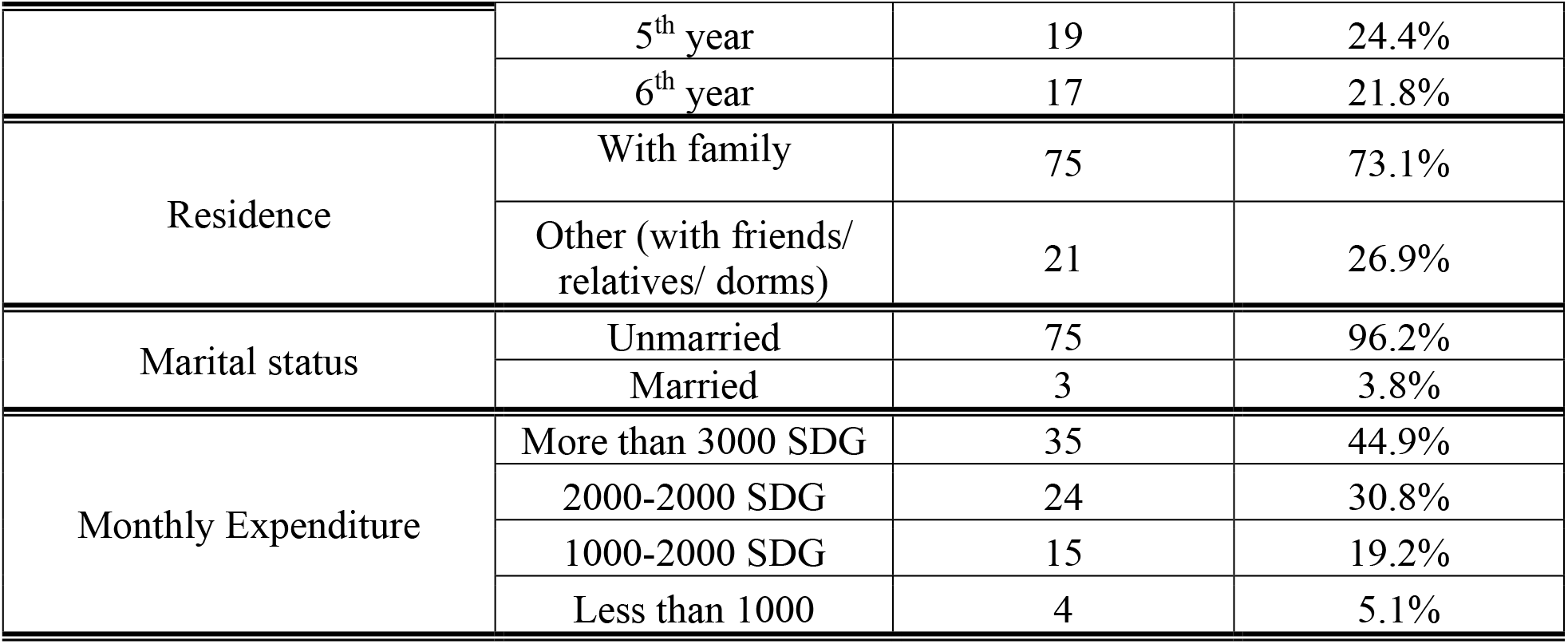
Summary of participants’ demographic information (N=78)

### Quality of Life Assessment (SF-12)

The SF-12 scores revealed that 42.3% of the students reported a poor quality of life. Of the eight subscales of HRQoL, participants scored highest on the physical functioning domain (Mean=73.08), while the lowest score was on the emotional role domain (Mean=28.21). The subscales that showed poor scores (>50) were Physical Role (RP), Social Functioning (SC), and Emotional Role (RE) [Table 2]. Subjects scored significantly lower on the mental component (MCS) (43.9 ± 19.4) than the physical component (PCS) (63.3 ± 23.2) (P > 0.01). A comparison between genders showed that female students reported a lower PCS score compared to male students (P=0.002) indicating a poorer physical quality of life among females. Other sociodemographic information showed no statistically significant correlation with the SF-12 components.

**Table 2:**
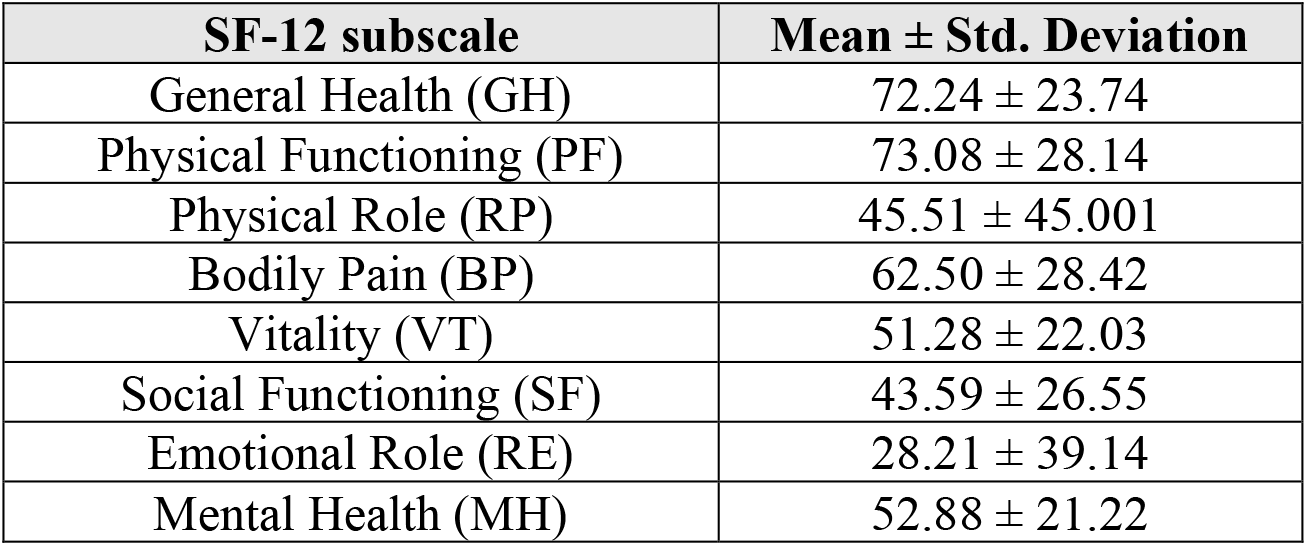
Participants’ scores on the eight domains of quality of life (N=78)

| **SF-12 subscale** | **Mean ± Std. Deviation** |
| --- | --- |
| General Health (GH) | 72.24 ± 23.74 |
| Physical Functioning (PF) | 73.08 ± 28.14 |
| Physical Role (RP) | 45.51 ± 45.001 |
| Bodily Pain (BP) | 62.50 ± 28.42 |
| Vitality (VT) | 51.28 ± 22.03 |
| Social Functioning (SF) | 43.59 ± 26.55 |
| Emotional Role (RE) | 28.21 ± 39.14 |
| Mental Health (MH) | 52.88 ± 21.22 |

### Psychiatric Comorbidities (HSCL-25)

The HSCL-25 scores indicated that psychiatric symptoms were prevalent among the participants. The mean score for the depression and anxiety subscales and the overall HSCL-25 score were all higher than the recommended cutoff point which is 1.75 (2.26, 2.11, and 2.20 respectively). 70.5% of subjects scored higher than the cutoff point in both subscales, while 71.8% scored higher than the cutoff point on the overall scale [Table 3]. Female gender showed a statistically significant correlation with HSCL-25 scores, with females reporting higher scores than males (P= 0.021 for the overall HSCL-25 score, and 0.045 and 0.011 for the depression and anxiety subscales respectively). Other sociodemographic information showed no statistically significant correlation with psychiatric symptoms.

**Table 3:** Mean and SD of the depression and anxiety subscales scores, and number and percentage of subjects who scored higher than the cutoff point (N=78)

|  | **Mean** | **Std.**<br>**Deviation** | **No. of Subjects**<br>**>1.75** | **% of**<br>**Subjects**<br>**>1.75** |
| --- | --- | --- | --- | --- |
| **HSCL**<br>**depression**<br>**subscale** | 2.26 | 0.69 | 55 | 70.5% |
| **HSCL anxiety**<br>**subscale** | 2.11 | 0.62 | 55 | 70.5% |
| **HSCL-25** | 2.20 | 0.63 | 56 | 71.8% |

## Discussion

This study aimed to assess the health-related quality of life and psychiatric comorbidities among medical students. The findings revealed a substantial impairment in the quality of life, particularly in the mental component, with 42.3% of students reporting poor quality of life. Additionally, psychiatric comorbidities, including anxiety and depression, were prevalent, affecting over 70% of the participants. Notably, female students reported poorer physical quality of life and higher rates of psychiatric symptoms than their male counterparts, consistent with previous research [16].

In line with existing literature, most participants in this study were female (74.4%), reflecting the higher prevalence of migraine in women as reported by Stovner et al. (2007) and Teixeira et al. (2012) [17,18]. The SF-12 scores revealed a significant disparity between the mental and physical components of quality of life, with the Mental Component Summary (MCS) score being notably lower than the Physical Component Summary (PCS) score. This finding contrasts with the finding from Monzon et al. (1998) where the physical domain was more affected, suggesting that the mental health burden in our population may be particularly severe [19].

Among the eight domains of quality of life, social functioning (SF) scored the lowest, again differing from findings by Monzon et el. (1998) as well as Terwindt et el. (2000) who reported lower scores in vitality (VT), general health (GH), and bodily pain (BP) [19, 20]. The gender disparity in quality of life, with females reporting worse physical health outcomes, is consistent with the study by Saif et al. (2017) that highlighted the greater impact of migraine on women [21].

The psychiatric assessment using the Hopkins Symptom Checklist-25 (HSCL-25) revealed high rates of anxiety and depression, both at 70.5%. These findings align with the existing previous research demonstrating a strong association between migraine and psychiatric comorbidities [22, 23, 24, 25]. The higher prevalence of psychiatric symptoms among female students is consistent with findings from Bensenor et el. (2003) showing that women are more vulnerable to developing anxiety and depression [26].

One of the strengths of this study is its focus on a specific population—medical students—who may be particularly vulnerable to the effects of migraine due to their stressful academic environment. The use of validated tools like the SF-12 and HSCL-25 adds rigor to the assessment of quality of life and psychiatric symptoms. However, there are limitations, including the cross-sectional design, small sample size, and reliance on self-reported data, which may introduce reporting bias. Additionally, the absence of a control group and the lack of adjustment for confounding factors such as headache severity and sleep disorders are significant limitations.

## Conclusion and Recommendations

In conclusion, our study highlights the significant impact of migraines on both the physical and mental health of medical students, with 42.3% reporting poor overall health-related quality of life. Most students reported high levels of anxiety and depression, with 71.8% suffering from emotional distress. Females were particularly affected, showing poorer quality of life and higher rates of psychiatric symptoms.

These findings emphasize the need for comprehensive management strategies that address both the physical and mental health aspects of migraine, particularly in high-stress populations such as medical students. Universities should consider implementing support programs that include regular mental health screenings, counseling services, and tailored medical management for migraine sufferers. Such initiatives could help mitigate the impact of migraine on students’ academic performance and overall well-being. Finally, future research should explore these areas further, ideally through longitudinal studies with larger, more diverse samples.

## Data Availability

All data produced in the present work are contained in the manuscript

